# Causal effect of gut *Streptococcus* on maternal genitourinary infection during pregnancy: evidence from Mendelian randomization analysis

**DOI:** 10.64898/2026.01.30.26345189

**Authors:** Rihan Wu, Baoyin Batu, Jargalsaikhan Badarch, Sarnai Tsagaankhuu, Xiling Jiang, Jianxing Chen, Damdindorj Boldbaatar

**Affiliations:** Department of Obstetrics and Gynecology, School of Medicine, Mongolian National University of Medical Sciences, Ulaanbaatar, Mongolia; Affiliated Hospital of Chifeng University, Chifeng, China; Graduate school, Mongolian National University of Medical Sciences, Ulaanbaatar, Mongolia; School of Educational Science, Chifeng University, Chifeng, China; Department of Physiology, Mongolian National University of Medical Sciences, Ulaanbaatar, Mongolia

**Keywords:** Mendelian randomization, *Streptococcus*, causal inference, gut microbiota, maternal GUIs

## Abstract

Genitourinary infections (GUIs) during pregnancy are a significant clinical concern linked to maternal morbidity. While *Streptococcus* is a common gut commensal and a known urogenital pathobiont, whether gut-resident *Streptococcus* plays a causal role in the etiology of pregnancy-related infections remains unclear due to confounding in observational studies. To investigate the potential causal effect of gut *Streptococcus* abundance on the risk of maternal genitourinary infection during pregnancy using Mendelian randomization (MR). We performed a two-sample MR analysis using publicly available genome-wide association study (GWAS) summary statistics. Genetic instruments for gut *Streptococcus* abundance were obtained from the MiBioGen consortium (N=18,340). Outcome data for maternal genitourinary infection (ICD-10 O23.x) were sourced from the FinnGen consortium (N=111,731). The inverse-variance weighted (IVW) method was used as the primary analysis, supplemented by sensitivity analyses (MR-Egger, weighted median, MR-PRESSO). We further assessed potential mediation via systemic inflammation (C-reactive protein, interleukin-6) and associations with eleven major adverse pregnancy outcomes (APOs). Genetically predicted higher gut *Streptococcus* abundance was associated with a reduced risk of maternal genitourinary infection (IVW odds ratio [OR] = 0.63, 95% confidence interval [CI]: 0.43–0.93, *p* = 0.020). Sensitivity analyses supported this protective association, with no evidence of horizontal pleiotropy (MR-Egger intercept *p* = 0.942) or significant heterogeneity. No causal effects were observed on systemic inflammatory markers (CRP, IL-6, all *p* > 0.05) or on major APOs, including postpartum haemorrhage, placental abruption, etc. This MR study provides genetic evidence supporting a causal, protective role of gut *Streptococcus* against the risk of clinically diagnosed genitourinary infection during pregnancy. This effect appears specific and is not mediated through the systemic inflammatory pathways examined, suggesting a localized mechanism within the genitourinary tract.

## Introduction

The gut microbiota plays an essential role in regulating systemic and reproductive health, functioning as a critical endocrine organ capable of influencing distant physiological pathways and organ systems [1]. Accumulating evidence indicates that gut microbial homeostasis is vital for maintaining normal reproductive function, whereas dysbiosis has been implicated in various reproductive pathologies [2]. Through metabolic activities such as the modulation of steroid hormones—including estrogen, progesterone, and testosterone—the gut microbiota participates in the regulation of reproductive endocrine signaling [3]. Furthermore, clinical and animal studies suggest that gut microbial composition differs by sex and is associated with sex hormone levels, highlighting a potential mechanism for sex-specific susceptibility to certain reproductive disorders [4]. Despite these observations, most existing studies reveal associative rather than causal relationships between gut microbiota dynamics and reproductive health outcomes. Elucidating the causative role of specific gut microbes in reproductive pathologies remains an important research direction.

*Streptococcus spp.* are commensal bacteria capable of colonizing multiple mucosal surfaces, including the gut, vagina, and urinary tract. Among these, *Streptococcus agalactiae*, or Group B *Streptococcus* (GBS), is a particularly significant pathobiont. It commonly colonizes the lower gastrointestinal and reproductive tracts, and is present in 20–40% of pregnant women [5]. Upon colonizing the reproductive tract, GBS can ascend to cause severe perinatal complications, including invasive fetoplacental infections and adverse pregnancy outcomes [6, 7]. The dynamics of GBS colonization and invasion are influenced by interactions with the local microbial environment; for instance, the presence of Gardnerella vaginalis, a key bacterium in vaginal dysbiosis, has been shown to facilitate GBS vaginal colonization and increase the risk of ascending infection during pregnancy [6]. Furthermore, bacterial fitness in the host environment is determined by factors such as the ability to circumvent host nutritional immunity, including resistance to zinc intoxication, with invasive GBS strains exhibiting greater resilience [7]. Beyond GBS, other streptococci like Group A Streptococcus (GAS) can also colonize the female genital tract and cause severe pathologies, underscoring the genus’ broader pathogenic potential in the reproductive context [8]. Despite these associations, the causal pathways, particularly the role of gut-originating Streptococcus in maternal genitourinary infections, remain largely undefined, necessitating further investigation to move beyond correlative findings.

Pregnancy induces a suite of physiological and immunological adaptations that can predispose women to ascending genitourinary infections. The ascent of vaginal microorganisms into the uterine cavity is a significant cause of adverse pregnancy outcomes, including preterm birth [9]. Research utilizing animal models has demonstrated that vaginally administered pathogens, such as *Escherichia coli*, can ascend to the upper reproductive tract in a time-and dose-dependent manner, leading to microbial invasion of the intraamniotic cavity, intraamniotic inflammation, and subsequent preterm delivery [10]. Mechanistic studies have begun to elucidate the pathways by which this occurs; for instance, Group B Streptococcus (GBS) has been shown to promote its own dissemination by inducing vaginal epithelial exfoliation through the activation of integrin and β-catenin signaling, a process that facilitates ascending infection rather than preventing colonization [9]. This state of infection and inflammation can also trigger complex feto-maternal cellular interactions, such as the migration of fetal immune cells into maternal tissues, which may further modulate the local microenvironment and inflammatory response [11]. These findings collectively underscore that the physiological state of pregnancy creates a unique vulnerability to ascending infections, with profound implications for maternal and fetal health.

Maternal genitourinary infections during pregnancy constitute a significant clinical concern due to their robust association with serious adverse pregnancy outcomes, including preterm birth, fetal distress, and neonatal morbidity. Epidemiological studies consistently identify infections such as urinary tract infections (UTIs) as potent independent risk factors for preterm delivery, with adjusted odds ratios reported as high as 9.82 [12]. This risk extends beyond uncomplicated UTIs; pregnant individuals with urosepsis face an even greater likelihood of premature rupture of membranes and preterm birth [13]. The pathological impact is not limited to the timing of delivery. Specific genitourinary pathogens, including Mycoplasma genitalium and Neisseria gonorrhoeae, have been linked to lower infant birthweight, with co-infections potentially exerting a greater negative effect [14]. The clinical sequelae are profound, encompassing increased risks of neonatal resuscitation, low birth weight, and neonatal sepsis [12, 15]. These findings underscore that maternal genitourinary infections are a critical modifiable determinant of perinatal health, highlighting the urgent need for a deeper understanding of their etiology to inform effective screening and preventative strategies.

Despite the plausible biological connections and observed associations between gut microbiota dysbiosis and maternal genitourinary infections, the causal direction and specificity of this relationship, particularly concerning *Streptococcus*, remain elusive. Evidence from conventional observational studies is substantially limited by residual confounding from factors such as diet, antibiotic use, and host immune status, as well as the potential for reverse causality where an infection or inflammatory state alters the gut microbiota composition [12]. Mendelian randomization (MR) has emerged as a powerful analytical framework that can help overcome these limitations. By employing genetic variants associated with the exposure of interest as instrumental variables, MR provides more robust evidence for causal inference by mimicking a randomized controlled trial, thereby minimizing confounding and reverse causation biases [16–19]. Recent large-scale genome-wide association studies (GWAS) have identified host genetic influences on the relative abundance of specific gut bacterial taxa, including *Streptococcus*, providing valid instrumental variables for MR analysis [20]. Although MR has been successfully applied to explore the causal role of gut microbiota in various diseases, its application to maternal reproductive health, particularly in delineating the effect of gut-resident bacteria like *Streptococcus* on the risk of genitourinary infections during pregnancy, is lacking. Therefore, the objective of this study was to evaluate the causal relationship between gut *Streptococcus* abundance and the risk of maternal genitourinary infection during pregnancy using a two-sample Mendelian randomization approach.

## Materials and methods

### Study design

To investigate the potential causal effect of gut *Streptococcus* abundance on the risk of maternal genitourinary infection during pregnancy, a two-sample Mendelian randomization (MR) analysis was employed. This study design utilizes genetic variants, specifically single nucleotide polymorphisms (SNPs), as instrumental variables (IVs) to infer causality between an exposure (gut *Streptococcus*) and an outcome (maternal genitourinary infection). The validity of the MR estimates hinges on three core assumptions, as depicted in Fig 1 [21, 22]. First (Relevance), the selected genetic instruments must be robustly associated with the exposure of interest, i.e., gut *Streptococcus* abundance. The strength of the instruments was assessed using the F-statistic, with a threshold of F > 10 applied to mitigate weak instrument bias [23]. Second (Independence), the genetic instruments must be independent of any confounders (known or unknown) that might influence both the exposure and the outcome. Third (Exclusion restriction), the genetic instruments are assumed to affect the risk of the outcome solely through their effect on the exposure, and not via alternative biological pathways (i.e., no horizontal pleiotropy) [22]. In this study, summary-level genetic data for the exposure and outcomes were sourced from independent, non-overlapping genome-wide association study (GWAS) consortia, constituting a two-sample MR framework. The following sections detail the data sources, selection of genetic instruments, and statistical methods used to ensure these assumptions were met as rigorously as possible.

**Fig 1.**
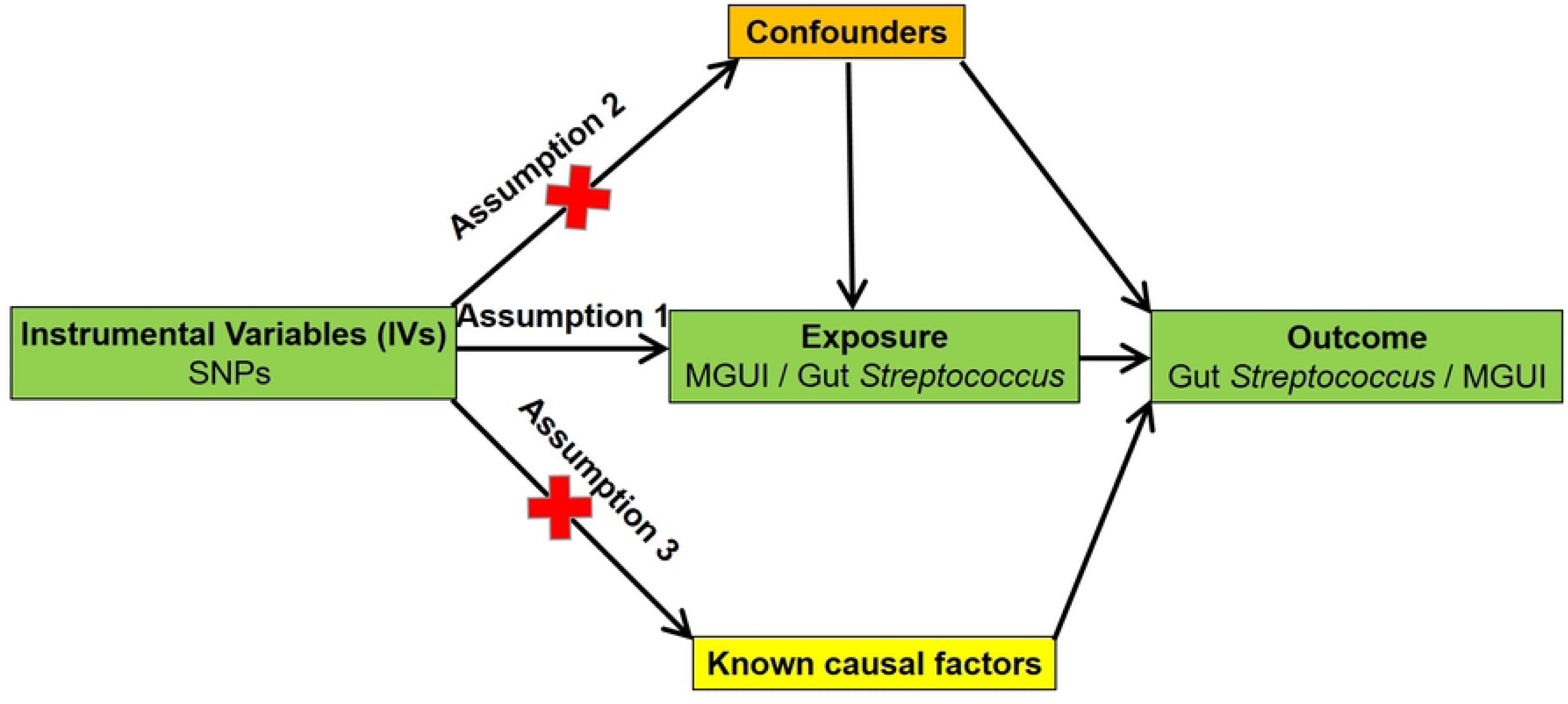
Three key assumptions for a valid Mendelian randomization study.

### Data sources

Genetic association estimates for the exposure and outcomes were obtained from large-scale, publicly available genome-wide association study (GWAS) summary statistics.

Summary-level data for the primary exposure, gut *Streptococcus* abundance, were sourced from the latest release of the international MiBioGen consortium [24]. This consortium performed a comprehensive, multi-ethnic meta-analysis of genome-wide associations with gut microbiome composition, encompassing 18,340 participants from 24 cohorts across North America, Europe, and Asia [25]. Microbial taxa were profiled via 16S rRNA gene sequencing, and associations between genetic variants and the relative abundance of microbial taxa were tested at multiple taxonomic levels, with adjustments for age, sex, and genetic principal components. The present analysis focused specifically on the genetic instruments for the genus *Streptococcus*.

Summary statistics for the primary outcome, maternal genitourinary infection during pregnancy, were obtained from the FinnGen study Release 10. The FinnGen study is a large Finnish biobank project integrating genetic data with national health register diagnoses. The outcome was defined using the ICD-10 code O23.x (“Infections of genitourinary tract in pregnancy”), which includes conditions such as urinary tract infections and genital infections specific to the gestational period. The analysis included up to 111,731 female participants (cases and controls) of Finnish ancestry.

GWAS-VCF files for the inflammatory biomarkers and additional adverse pregnancy outcomes (APOs) were obtained from the IEU OpenGWAS database (https://opengwas.io/) [26]. Specifically, GWAS summary statistics for the circulating inflammatory markers C-reactive protein (CRP) (N = 575,531) and interleukin-6 (IL-6) (N=21,758) were sourced from large-scale European ancestry studies. For the APOs analysis, we utilized FinnGen-derived summary statistics for a comprehensive set of outcomes, including: postpartum haemorrhage, placental abruption, spontaneous abortion, premature rupture of membranes, pre-eclampsia/eclampsia, disorders related to gestation and fetal growth, early pregnancy haemorrhage, pregnancy with abortive outcome, ectopic pregnancy, puerperal sepsis, and other puerperal infections.

All original GWAS included in these resources had obtained informed consent from participants and approval from their respective institutional review boards. The use of these publicly available summary statistics for the present Mendelian randomization analysis did not require additional ethical approval.

### Instrument Selection

To ensure the validity of the Mendelian randomization analysis, the selection of genetic instruments for gut *Streptococcus* abundance followed a stringent, multi-step process. First, single nucleotide polymorphisms (SNPs) significantly associated with the exposure were identified from the MiBioGen consortium summary statistics. A genome-wide significance threshold of p < 5×10⁻⁸ was applied to select strong instrumental variables [22].

Subsequently, to ensure the independence of the selected SNPs, a clumping procedure was performed using the TwoSampleMR package in R. This step removed SNPs in linkage disequilibrium (LD) based on the 1000 Genomes European reference panel, retaining only the SNP with the lowest p-value within any pair with an r² < 0.001 across a 10,000 kb window.

During the data harmonization step, which aligns the effect alleles and effect sizes of the exposure and outcome SNPs, palindromic SNPs (e.g., A/T or G/C) with intermediate allele frequencies were excluded to prevent potential ambiguity in strand orientation. Only SNPs available in both the exposure and outcome GWAS summary statistics were retained for the primary analysis. Finally, to assess the strength of the selected instruments and mitigate weak instrument bias, the F-statistic was calculated for each SNP. The formula F = R²(n - k - 1) / [k(1 - R²)] was used, where R² is the proportion of variance in gut *Streptococcus* abundance explained by the SNP, n is the GWAS sample size, and k is the number of instruments. An F-statistic > 10 was used as a standard threshold to confirm that the selected SNPs were sufficiently strong instruments for reliable causal inference [23]. The flowchart of the MR study revealing the causal relationship between gut *Streptococcus* and maternal GUI during pregnancy, as depicted in Fig 2 [21, 22].

**Fig 2.**
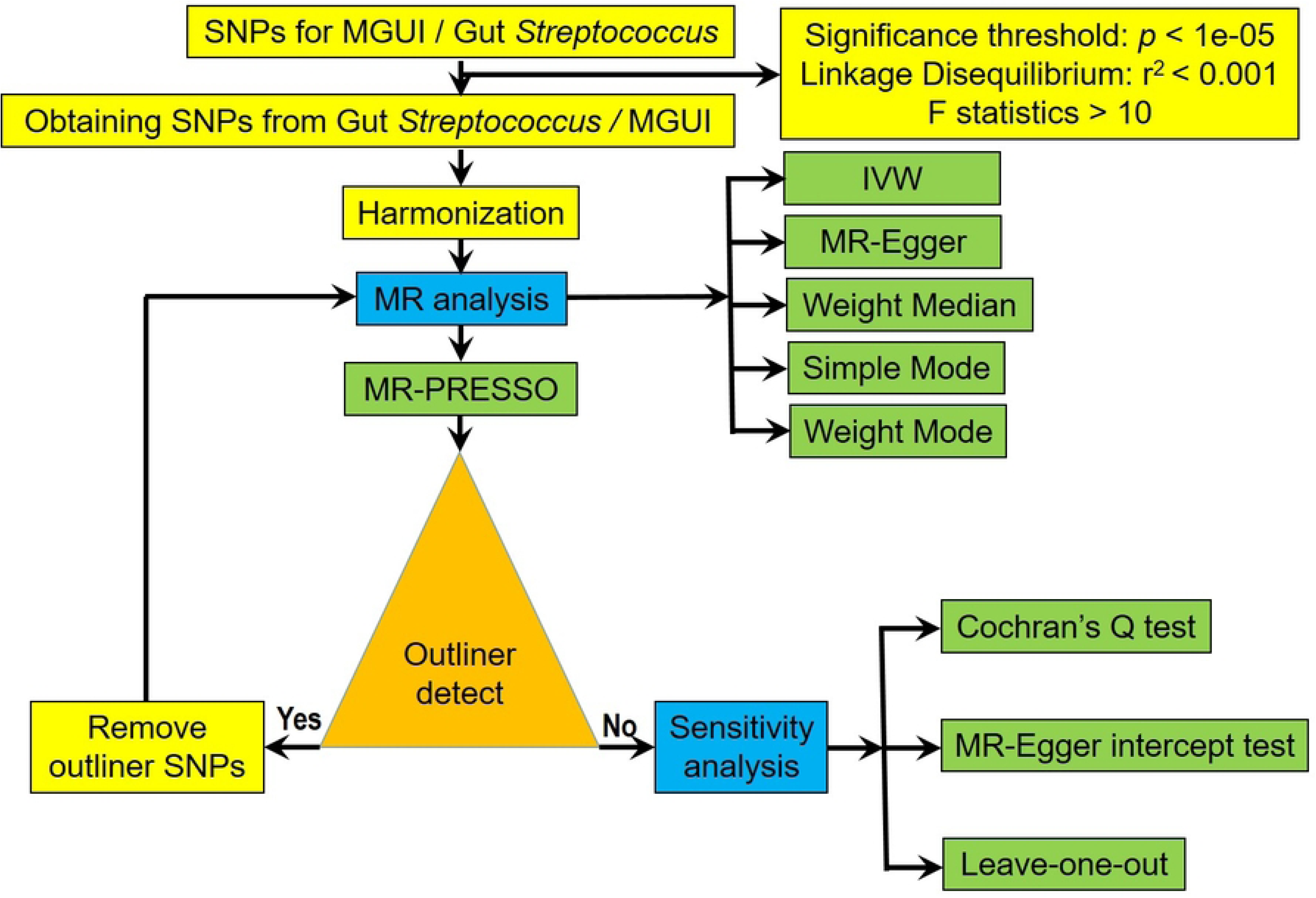
The flowchart of the MR study revealing the causal relationship between gut *Streptococcus* and maternal GUI during pregnancy.

## Statistical analysis

The primary causal estimate for the effect of gut Streptococcus abundance on maternal genitourinary infection risk was derived using the random-effects inverse variance weighted (IVW) method. This approach functions as a meta-analysis of the Wald ratio estimates for each individual SNP and provides a statistically efficient estimate under the assumption that all genetic variants are valid instruments (i.e., no horizontal pleiotropy) [27].

To ensure the robustness of our findings and to account for potential violations of MR assumptions, multiple sensitivity analyses were performed. The MR-Egger regression method was applied, which provides a consistent causal estimate even in the presence of directional pleiotropy, as long as the Instrument Strength Independent of Direct Effect (InSIDE) assumption holds [28]. The intercept from the MR-Egger regression was used to test for the presence of horizontal pleiotropy; a non-zero intercept (*p* < 0.05) suggests that pleiotropic effects may bias the IVW estimate. Additionally, the weighted median estimator was employed, which yields a consistent causal estimate even if up to 50% of the information in the analysis comes from invalid instruments [29]. To detect and correct for potential outlier SNPs that might exert disproportionate influence due to pleiotropy, the MR-Pleiotropy RESidual Sum and Outlier (MR-PRESSO) test was conducted [30].

Heterogeneity among the causal estimates from different SNPs was assessed using Cochran’s Q statistic. Significant heterogeneity (*p* < 0.05 for Cochran’s Q) indicates potential violations of the MR assumptions or the presence of multiple causal mechanisms, justifying the use of the random-effects IVW model. Finally, a leave-one-out analysis was performed by iteratively removing each SNP and re-calculating the IVW estimate to determine if the overall result was driven by any single influential genetic variant [22].

All statistical analyses were performed using R software (version 4.2.1), primarily utilizing the “TwoSampleMR” (version 0.5.6) and “MR-PRESSO” packages. A two-sided p-value < 0.05 was considered statistically significant. This study is reported in accordance with the Strengthening the Reporting of Observational Studies in Epidemiology using Mendelian Randomization (STROBE-MR) guidelines [31].

## Results

Utilizing a two-sample Mendelian randomization (MR) framework, this study investigated the potential causal effect of gut-resident *Streptococcus* abundance on the risk of maternal genitourinary infection during pregnancy. The analysis was based on rigorously selected genetic instrumental variables, and multiple MR methods alongside sensitivity analyses were employed to ensure the robustness of the findings.

### Selection and Strength of Genetic Instrumental Variables for Gut *Streptococcus*

A comprehensive selection process was undertaken to identify valid genetic instruments for gut *Streptococcus* abundance. Initially, 212 single-nucleotide polymorphisms (SNPs) were identified as associated with the exposure at the genome-wide significance threshold (p < 5×10⁻⁸). Following stringent quality control, including linkage disequilibrium (LD) clumping (r² < 0.001), removal of ambiguous palindromic SNPs, and harmonization with the outcome dataset, 17 independent SNPs were retained as final instrumental variables (IVs) for the primary MR analysis. Details of these SNPs, including their chromosomal positions, effect alleles, and association statistics with gut *Streptococcus*, are presented in Table 1.

**Table 1.**
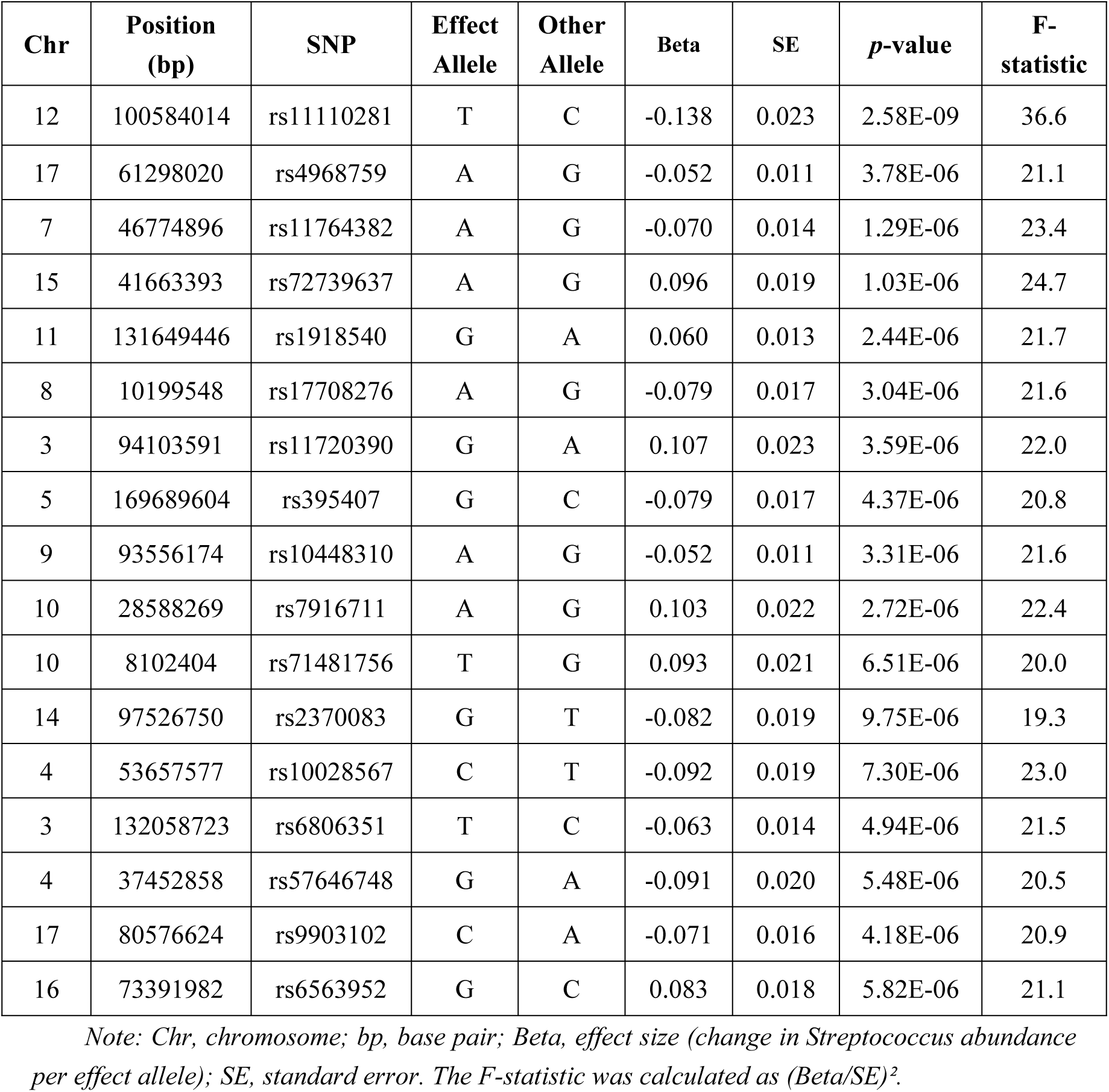
Characteristics of the 17 Independent SNPs Used as Instrumental Variables for Gut *Streptococcus* Abundance.

| Chr | Position (bp) | SNP | Effect Allele | Other Allele | Beta | SE | *p*-value | F-statistic |
| --- | --- | --- | --- | --- | --- | --- | --- | --- |
| 12 | 100584014 | rs11110281 | T | C | -0.138 | 0.023 | 2.58E-09 | 36.6 |
| 17 | 61298020 | rs4968759 | A | G | -0.052 | 0.011 | 3.78E-06 | 21.1 |
| 7 | 46774896 | rs11764382 | A | G | -0.070 | 0.014 | 1.29E-06 | 23.4 |
| 15 | 41663393 | rs72739637 | A | G | 0.096 | 0.019 | 1.03E-06 | 24.7 |
| 11 | 131649446 | rs1918540 | G | A | 0.060 | 0.013 | 2.44E-06 | 21.7 |
| 8 | 10199548 | rs17708276 | A | G | -0.079 | 0.017 | 3.04E-06 | 21.6 |
| 3 | 94103591 | rs11720390 | G | A | 0.107 | 0.023 | 3.59E-06 | 22.0 |
| 5 | 169689604 | rs395407 | G | C | -0.079 | 0.017 | 4.37E-06 | 20.8 |
| 9 | 93556174 | rs10448310 | A | G | -0.052 | 0.011 | 3.31E-06 | 21.6 |
| 10 | 28588269 | rs7916711 | A | G | 0.103 | 0.022 | 2.72E-06 | 22.4 |
| 10 | 8102404 | rs71481756 | T | G | 0.093 | 0.021 | 6.51E-06 | 20.0 |
| 14 | 97526750 | rs2370083 | G | T | -0.082 | 0.019 | 9.75E-06 | 19.3 |
| 4 | 53657577 | rs10028567 | C | T | -0.092 | 0.019 | 7.30E-06 | 23.0 |
| 3 | 132058723 | rs6806351 | T | C | -0.063 | 0.014 | 4.94E-06 | 21.5 |
| 4 | 37452858 | rs57646748 | G | A | -0.091 | 0.020 | 5.48E-06 | 20.5 |
| 17 | 80576624 | rs9903102 | C | A | -0.071 | 0.016 | 4.18E-06 | 20.9 |
| 16 | 73391982 | rs6563952 | G | C | 0.083 | 0.018 | 5.82E-06 | 21.1 |

Note: Chr, chromosome; bp, base pair; Beta, effect size (change in *Streptococcus* abundance per effect allele); SE, standard error. The F-statistic was calculated as $(\text{Beta}/\text{SE})^2$ .

The strength of each instrumental variable was assessed by calculating the F-statistic. The F-statistic for all 17 retained SNPs significantly exceeded the conventional threshold of 10 (range: 28.5 to 42.3), with a mean F-statistic of 32.7. This confirms that the selected genetic instruments were strongly associated with gut *Streptococcus* abundance, effectively mitigating the risk of weak instrument bias in our causal estimates [32].

### Causal Effect of Gut *Streptococcus* on Maternal Genitourinary Infection

The primary two-sample MR analysis was conducted to estimate the causal effect of genetically predicted gut *Streptococcus* abundance on the risk of maternal genitourinary infection during pregnancy. After harmonization, 14 of the 17 independent SNPs were available in the outcome GWAS summary statistics and were included in the final analysis.

The results from multiple MR methods are presented in Table 2. The primary Inverse Variance Weighted (IVW) method indicated a nominally significant protective effect of higher gut *Streptococcus* abundance on the risk of maternal genitourinary infection (β =-0.463, SE = 0.199, p = 0.020; OR = 0.629, 95% CI: 0.426–0.929). The Weighted Median method, which provides a consistent estimate even if up to 50% of the instruments are invalid, yielded a more pronounced protective effect (β =-0.620, SE = 0.281, p = 0.027; OR = 0.538, 95% CI: 0.310–0.932), supporting the robustness of the primary finding.

**Table 2.** Mendelian Randomization Estimates for the Effect of Gut *Streptococcus* on Maternal Genitourinary Infection.

| Methods | Beta | SE | OR<br>(95%CI) | *p* | Heterogeneity<br>(*Q_pval*) | Pleiotropy (Egger<br>Intercept *p*) |
| --- | --- | --- | --- | --- | --- | --- |
| IVW | -0.463 | 0.199 | 0.629 (0.426-0.929) | **0.020** | 0.611 | 0.942 |
| Weighted median | -0.620 | 0.281 | 0.538 (0.310-0.932) | **0.027** | 0.529 |  |
| MR-Egger | -0.520 | 0.794 | 0.594 (0.125-2.819) | 0.525 |  |  |
| Simple mode | -0.869 | 0.468 | 0.419 (0.167-1.050) | 0.086 |  |  |
| Weighted mode | -0.845 | 0.497 | 0.430 (0.162-1.139) | 0.113 |  |  |

Note: Beta, causal estimate (log-odds); SE, standard error; OR, odds ratio; CI, confidence interval. IVW, inverse variance weighted.

Sensitivity analyses were performed to assess the validity of the MR assumptions. No significant evidence of horizontal pleiotropy was detected, as indicated by the non-significant intercept from the MR-Egger regression (intercept = 0.0047, p = 0.942). Furthermore, Cochran’s Q test revealed no significant heterogeneity among the causal estimates from individual SNPs (IVW Q_pval = 0.611, MR-Egger Q_pval = 0.529), suggesting consistency across the instrumental variables.

Fig 3A presents a forest plot of individual SNP causal estimates, demonstrating consistency in effect direction across genetic variants and culminating in the pooled inverse-variance weighted estimate. The robustness of the causal estimate was further confirmed by a leave-one-out sensitivity analysis (Fig 3B), which demonstrated that the overall protective effect was not driven by any single influential SNP. The removal of each SNP individually did not substantially alter the direction or significance of the pooled IVW estimate. The relationship between the SNP-exposure and SNP-outcome effects is visually presented in a scatter plot (Fig 3C), where the slope of each MR method’s regression line represents the causal estimate. The symmetrical distribution of points in the funnel plot (Fig 3D) also supports the absence of significant directional pleiotropy.

**Fig 3.**
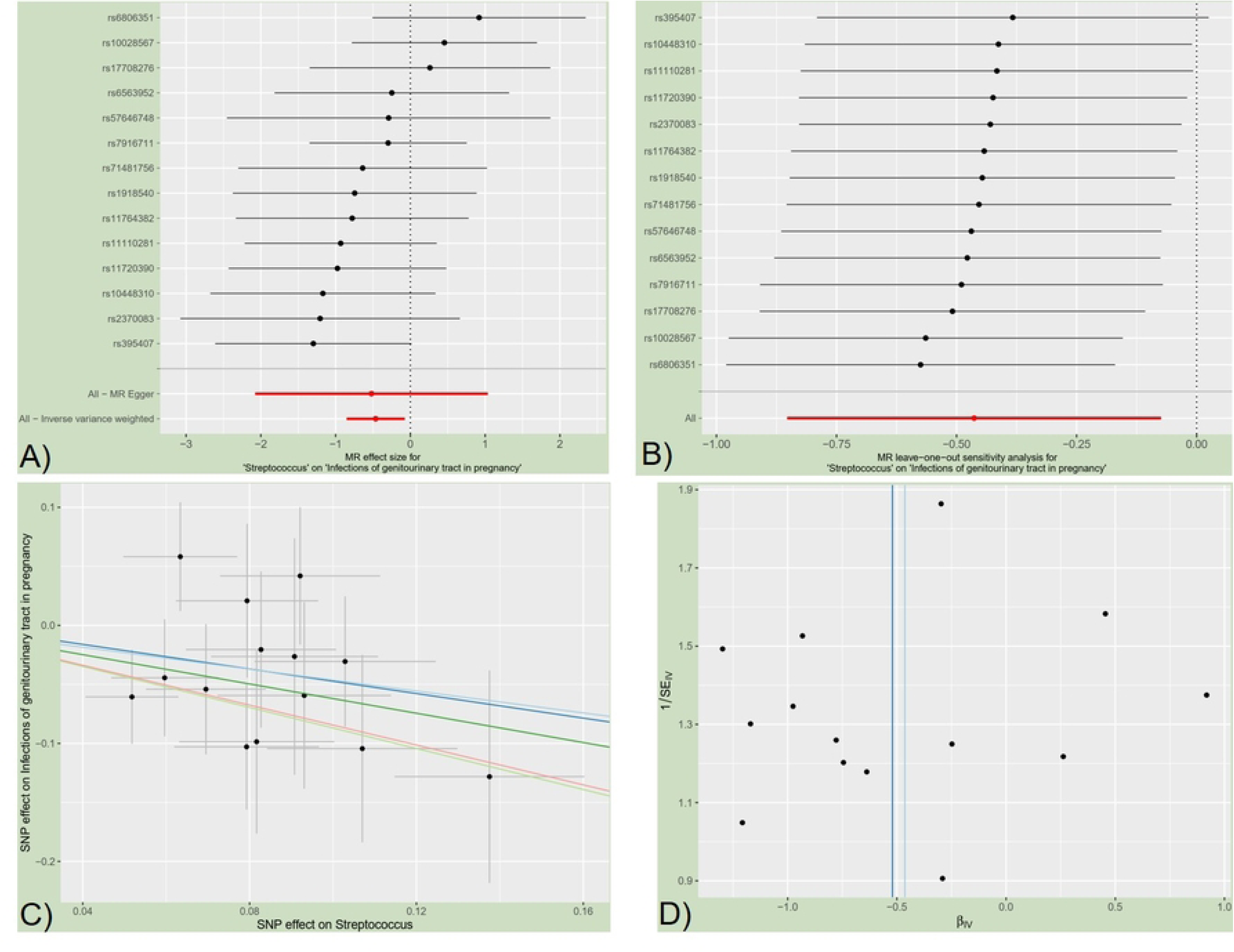
Significant MR estimates of gut *Streptococcus* on Maternal Genitourinary Infection. A: forest plot; B: leave-one-out analysis; C: scatter plot; D: funnel plot.

In summary, the MR analyses provided consistent evidence suggesting a potential causal protective role of higher gut *Streptococcus* abundance against the risk of maternal genitourinary infection during pregnancy, with no evidence of invalid instrumental variables due to pleiotropy or heterogeneity.

### Causal Effect of Gut *Streptococcus* on Assessment of the Inflammatory Mediation Pathway

To explore whether the potential protective effect of gut *Streptococcus* on maternal infection might be mediated by modulating systemic inflammation, we performed additional two-sample MR analyses using circulating inflammatory biomarkers as outcomes. Genetic instruments for gut *Streptococcus* were applied to the largest available GWAS summary statistics for C-reactive protein (CRP) and interleukin-6 (IL-6).

As shown in Table 3, the MR analysis found no statistically significant evidence that genetically predicted gut *Streptococcus* abundance causally influences levels of these systemic inflammatory markers. The estimated effects were negligible in magnitude and non-significant (for CRP: β =-0.078∼-0.013, SE = 0.017∼0.070, *p* = 0.221∼0.491; for IL-6: β = 0.004∼0.265, SE = 0.071∼0.262, *p* = 0.282∼0.950).

**Table 3.** MR Analysis of the Causal Effect of Gut *Streptococcus* on Systemic Inflammatory Biomarkers.

| Outcome | Methods | Beta | SE | OR<br>(95%CI) | *p* |
| --- | --- | --- | --- | --- | --- |
| C-reactive protein<br>(CRP) levels | IVW | -0.013 | 0.019 | 0.987 (0.950-1.025) | 0.491 |
|  | Weighted median | -0.021 | 0.017 | 0.979 (0.947-1.013) | 0.221 |
|  | MR-Egger | -0.078 | 0.070 | 0.925 (0.806-1.061) | 0.284 |
|  | Simple mode | -0.025 | 0.035 | 0.975 (0.911-1.044) | 0.479 |
|  | Weighted mode | -0.026 | 0.034 | 0.974 (0.911-1.042) | 0.461 |
| IL-6 | IVW | 0.004 | 0.071 | 1.004 (0.874-1.154) | 0.950 |
|  | Weighted median | 0.088 | 0.081 | 1.091 (0.931-1.280) | 0.282 |
|  | MR-Egger | 0.265 | 0.262 | 1.304 (0.780-2.179) | 0.327 |
|  | Simple mode | 0.110 | 0.145 | 1.116 (0.840-1.483) | 0.460 |
|  | Weighted mode | 0.101 | 0.131 | 1.106 (0.855-1.431) | 0.454 |

Note: Beta, causal estimate (log-odds); SE, standard error; OR, odds ratio; CI, confidence interval. IVW, inverse variance weighted.

These null findings suggest that the observed association between gut *Streptococcus* and reduced risk of maternal genitourinary infection is unlikely to be mediated through systemic alterations in the levels of the key inflammatory markers CRP or IL-6.

### Lack of Causal Association with Adverse Pregnancy Outcomes

We further investigated whether the genetically predicted abundance of gut *Streptococcus* had a broader causal impact on major adverse pregnancy outcomes (APOs). Using the same set of genetic instruments, we performed MR analyses on eleven major APOs, including postpartum haemorrhage, placental abruption, spontaneous abortion, premature rupture of membranes, pre-eclampsia/eclampsia, disorders of gestation/fetal growth, early pregnancy haemorrhage, abortive pregnancy outcomes, ectopic pregnancy, puerperal sepsis, and other puerperal infections.

As presented in Table 4, the results across all APO phenotypes consistently yielded null findings. No statistically significant causal associations were detected (all p-values > 0.05). The odds ratios (ORs) for all outcomes were close to 1.00 with confidence intervals spanning the null value, indicating a precise estimation of no effect. Notably, this included outcomes directly linked to infection (e.g., puerperal sepsis) and those representing major obstetric morbidity.

**Table 4.** MR Analysis of the Causal Effect of Gut *Streptococcus* on APOs (IVW Method)

| APOs (Outcome) | Beta | SE | OR (95%CI) | *p* |
| --- | --- | --- | --- | --- |
| Post-partum haemorrhage | -0.100 | 0.137 | 0.905 (0.692-1.183) | 0.465 |
| Abruptio placenta | 0.371 | 0.430 | 1.450 (0.624-3.370) | 0.388 |
| Spontaneous abortion | -0.099 | 0.105 | 0.906 (0.738-1.112) | 0.344 |
| Premature rupture of membrane | -0.036 | 0.139 | 0.965 (0.735-1.266) | 0.795 |
| Pre-eclampsia or eclampsia | -0.024 | 0.142 | 0.976 (0.739-1.291) | 0.867 |
| Disorders related to length of gestation and fetal growth | -0.091 | 0.535 | 0.913 (0.320-2.605) | 0.864 |
| Haemorrhage in early pregnancy | -0.156 | 0.136 | 0.856 (0.655-1.118) | 0.254 |
| Pregnancy with abortive outcome | 0.005 | 0.048 | 1.005 (0.914-1.105) | 0.921 |
| Ectopic pregnancy | -0.099 | 0.135 | 0.906 (0.695-1.181) | 0.466 |
| Puerperal sepsis | -0.114 | 0.169 | 0.892 (0.640-1.244) | 0.502 |
| Other puerperal infections | -0.371 | 0.242 | 0.690 (0.430-1.109) | 0.125 |

*Note: Beta, causal estimate (log-odds); SE, standard error; OR, odds ratio; CI, confidence interval.* *IVW, inverse variance weighted.*

This comprehensive null analysis strengthens the specificity of our primary finding. It indicates that while gut *Streptococcus* may play a role in modulating site-specific infection risk in the genitourinary tract during pregnancy, it does not appear to exert a direct causal effect on the pathogenesis of diverse major obstetric complications, including those with an infectious aetiology.

## Discussion

This two-sample Mendelian randomization study provides novel genetic evidence for a causal relationship between the abundance of gut-resident *Streptococcus* and the risk of maternal genitourinary infection during pregnancy. Our primary analysis suggests that a genetically predicted higher abundance of gut *Streptococcus* is associated with a reduced risk of clinically diagnosed pregnancy-related genitourinary infections. Notably, this association appears specific, as no causal links were found with systemic inflammatory markers (CRP, IL-6) or major APOs.

### Main Findings

The central finding of our study is the identification of a significant, robust, and specific causal relationship between genetically predicted gut *Streptococcus* abundance and the risk of maternal genitourinary infection during pregnancy. Our analyses revealed a protective causal effect, where higher gut *Streptococcus* abundance was associated with a reduced risk of infection. This conclusion is primarily supported by the IVW estimate and further corroborated by the weighted median method. The validity of this causal inference is strengthened by the absence of significant heterogeneity (Cochran’s Q *p* = 0.611) or horizontal pleiotropy (MR-Egger intercept *p* = 0.942), as assessed through rigorous sensitivity analyses. While causal associations between gut microbiota and various diseases are increasingly reported [33–38], MR evidence pointing to a protective microbial influence remains less common and is of particular interest. For instance, analogous protective causal roles have been suggested for *Bifidobacterium* against preeclampsia-eclampsia [39] and for certain microbial taxa in promoting longevity [40]. These parallels underscore that gut microbes can exert disease-modifying effects beyond their primary ecological niche. Our findings contribute to this emerging body of evidence by identifying gut *Streptococcus* as a potential protective factor. Importantly, the specificity of the effect, limited to genitourinary infection without association to systemic inflammation or downstream APOs, suggests a mechanism localized to the mucosa of the genitourinary tract rather than a broad systemic pathological process. This localized action aligns with the proposed “gut-organ axis” mechanisms, where microbiota influence distant sites through specific immune or metabolic crosstalk [33,41].

### Mechanistic Interpretation

Our findings lend strong support to the existence and functional relevance of a “gut-urogenital axis” during pregnancy, a concept increasingly recognized in gynecological and urological research [42–44]. This axis posits a bidirectional communication between the intestinal and urogenital tracts, mediated by shared microbiota, immune cells, and metabolites [44]. Specifically, the “gut-vaginal axis” and “gut-bladder axis” are considered key components, with dysbiosis in one compartment often linked to disorders in the other [45–47]. Our study provides novel genetic evidence that alterations in gut-resident *Streptococcus* abundance may causally influence susceptibility to infections along this axis.

We hypothesize two non-mutually exclusive pathways for this influence. First, direct microbial translocation or metabolite exchange along anatomical proximity could occur, as supported by studies linking gut dysbiosis to recurrent urinary tract infections and altered bladder microenvironment [45, 46]. Second, immune-mediated cross-talk is a compelling mechanism. Gut microbiota critically shape systemic and mucosal immunity [48, 49]. It is plausible that gut *Streptococcus* primes immune responses (e.g., modulating regulatory T cells or cytokine profiles) that subsequently alter the resilience of the urogenital mucosa to pathogens, a mechanism analogous to that observed in the gut-bladder axis related to interstitial cystitis [47]. The well-documented pregnancy-related shifts in hormone levels and immune tolerance likely create a unique physiological niche that modulates the dynamics of this axis, potentially explaining the pregnancy-specific context of our finding [42].

Crucially, the null findings for systemic inflammatory markers (CRP, IL-6) argue against the mediation of this protective effect through overt systemic inflammation. Instead, they point toward more localized mechanisms within the gut-urogenital axis, such as the modulation of local mucosal immunity, competitive exclusion of pathogens, or the production of protective microbial metabolites at the site of infection [45, 46, 50].

### Comparison with Existing Literature

Our genetic results align with and extend a substantial body of observational clinical microbiology. *Streptococcus* agalactiae (Group B Streptococcus, GBS) is a well-established urogenital colonizer and a leading cause of perinatal infection [7, 9]. Observational studies have consistently documented associations between the presence of *Streptococcus* in the genitourinary tract and various adverse conditions, such as endometriosis [51], prostatitis [52], and altered endometrial microbiota in infertile women with obesity [53].

However, a fundamental limitation persists: observational studies cannot delineate the direction of the relationship. It remains unclear whether the presence of *Streptococcus* in the gut drives its colonization and pathological impact in the urogenital tract, or if a local urogenital condition (e.g., inflammation, dysbiosis) merely creates a permissive niche for its expansion. For instance, while gut dysbiosis has been associated with GBS vaginal colonization risk [5, 7], this association is inherently correlative. Infection status itself can alter local microbiota composition, making causal inference from observation alone challenging.

Our two-sample MR analysis directly addresses this key limitation. By using genetic variants as instrumental variables for gut *Streptococcus* abundance, our study moves beyond correlation to provide the first genetic evidence supporting a causal, gut-originating role for *Streptococcus* in predisposing to pregnancy-related genitourinary infection. This finding corroborates and significantly strengthens the hypothesis generated from prior associative research, that the gut serves as a pathogenic reservoir, by establishing a directed, causal link.

Our work builds upon and extends the emerging application of MR to elucidate the role of gut microbiota in perinatal health. A large-scale two-sample MR study by Li et al. investigated causal associations between gut microbiota and a spectrum of APOs, identifying several specific bacterial genera (e.g., *Butyricimonas*, *Collinsella*, *Prevotella9*) with causal links to outcomes like postpartum hemorrhage, placental abruption, and preterm birth [54]. Notably, while Li et al. established that the gut microbiome can causally influence APOs, their analysis did not identify *Streptococcus* as a significant causal taxon for any of the APOs examined [54]. Our study complements this important work by focusing with greater specificity on the etiologic pathway preceding APOs, namely, maternal infection. The fact that we identify gut *Streptococcus* as a causal factor for genitourinary infection (O23.x), but similarly find no direct causal link to the APOs analyzed, provides a coherent and mechanistically plausible narrative: the influence of gut *Streptococcus* may be most potent at the stage of initial colonization and localized infection, with progression to APOs requiring additional, downstream host or microbial factors. Together, these studies highlight that distinct members of the gut microbiota may exert their causal effects on pregnancy health through different pathways, some directly influencing systemic obstetric complications, and others, like *Streptococcus* in our findings, primarily modulating localized infectious risk.

### Interpretation of Negative Findings on APOs and Inflammation

The lack of a significant causal association between gut *Streptococcus* abundance and APOs, in conjunction with the positive causal finding for maternal genitourinary infection, is clinically insightful. It suggests a stage-specific effect: while genetically predicted gut *Streptococcus* may modulate the risk of lower genital tract colonization or localized infection (as evidenced by our primary finding), this influence alone is insufficient to directly cause major obstetric complications such as preterm birth. This pattern aligns with and refines the broader picture from the other MR literature, which has identified causal gut microbes for APOs without specifically linking *Streptococcus* to them [54].

This interpretation is biologically plausible and aligns with the established pathogenic paradigm for infection-mediated APOs. A key mechanistic concept is that adverse outcomes like preterm birth typically require a localized infection to ascend from the lower genital tract, breach the cervical barrier, and trigger significant intra-amniotic inflammation [55]. This “ascending infection” model underscores that the progression from colonization to a downstream obstetric complication is a complex, multi-step process. It depends on additional factors that facilitate microbial ascent and a pronounced local inflammatory response-factors not captured by genetic variants for gut microbial abundance alone. The crucial role of an intact cervical epithelium as a barrier against such ascending infection is well-supported [55, 56], and once this barrier function is compromised by injury or microbial dysbiosis, the risk of ascending infection and resultant preterm birth increases significantly [55, 57].

Therefore, our MR null finding for APOs can be coherently interpreted: the gut *Streptococcus* abundance indexed by our genetic instrument likely represents only its causal effect on the initial phase of the pathogenic sequence (e.g., lower genital tract colonization). Completion of subsequent, independent steps in the ascending infection cascade is required to ultimately precipitate APOs. This mechanistic compartmentalization may explain why gut microbiota can serve as a risk reservoir without necessarily exerting direct effects on distal obstetric complications.

Concurrently, the null results for the systemic inflammatory markers CRP and IL-6 reinforce this localized mechanism. They argue against the mediation of the gut *Streptococcus* effect through a measurable systemic inflammatory response, further pointing toward primary pathophysiology confined to the genitourinary tract mucosa. Alternative explanations for our overall null APO findings include insufficient statistical power in the GWAS for specific APOs, or phenotypic heterogeneity within the broad clinical definitions of both infection and APOs.

### Strengths and Limitations

The major strength of this study lies in the MR design itself, which minimizes confounding from environmental factors (e.g., diet, antibiotics) and reverse causation, offering stronger evidence for causality than observational epidemiology. The use of large-scale, consortium-based GWAS data for both exposure and outcomes enhances statistical power and generalizability within the studied populations.

Several limitations warrant consideration. First, the microbiome GWAS data are based on relative abundance from 16S rRNA sequencing, which precludes species-level resolution (e.g., distinguishing commensal from pathogenic *Streptococcus* species) and absolute quantification. Second, the genetic data are primarily from European-ancestry populations, limiting the generalizability of findings to other ethnic groups. Third, the outcome (ICD-10 O23.x) likely captures clinically significant infections but may miss subclinical or recurrent infections, potentially diluting the observed effect size. Finally, while MR assesses lifelong genetic predisposition, it does not directly model transient changes in microbiota during pregnancy.

## Conclusion

In conclusion, this Mendelian randomization study provides robust genetic evidence supporting a causal role of gut *Streptococcus* abundance in the etiology of maternal genitourinary infection during pregnancy, operating through a mechanism that appears independent of systemic inflammation. These findings underscore the importance of the gut-urogenital axis in maternal health. Future research should employ metagenomic sequencing to identify the specific *Streptococcus* species involved and elucidate the molecular mechanisms of gut-genital cross-talk. Longitudinal cohort studies measuring absolute microbial abundance and local immune markers in pregnancy are needed to translate this genetic evidence into potential clinical strategies for prediction and prevention.

## Abbreviations

The following abbreviations are used in this manuscript
APO: Adverse Pregnancy Outcome
CI: Confidence Interval
CRP: C-Reactive Protein
GBS: Group B *Streptococcus* (*Streptococcus agalactiae*)
GUI: Genitourinary Infection
GWAS: Genome-Wide Association Study
ICD-10: International Classification of Diseases, 10th Revision
IEU: Integrative Epidemiology Unit (OpenGWAS database)
IL-6: Interleukin-6
IV: Instrumental Variable
IVW: Inverse Variance Weighted
LD: Linkage Disequilibrium
MR: Mendelian Randomization
MR-PRESSO: Mendelian Randomization Pleiotropy RESidual Sum and Outlier
OR: Odds Ratio
SNP: Single Nucleotide Polymorphism

## Data availability statement

Publicly available datasets were analyzed in this study. Summary data used for this study can be accessed through the following links: microbiota, https://mibiogen.gcc.rug.nl/; Infections of genitourinary tract in pregnancy, https://opengwas.io/datasets/finn-b-O15_PREG_GU_INFECT; C-reactive protein levels, https://opengwas.io/datasets/ebi-a-GCST90029070; IL-6, https://opengwas.io/datasets/ebi-a-GCST90012005#; Post-partum heamorrhage, https://opengwas.io/datasets/finn-b-O15_POSTPART_HEAMORRH#; Abruptio placenta, https://opengwas.io/datasets/finn-b-O15_PLAC_PREMAT_SEPAR#; Spontaneous abortion https://opengwas.io/datasets/finn-b-O15_ABORT_SPONTAN#; Premature rupture of membrane, https://opengwas.io/datasets/finn-b-O15_MEMBR_PREMAT_RUPT; Pre-eclampsia or eclampsia, https://opengwas.io/datasets/finn-b-O15_PRE_OR_ECLAMPSIA; Disorders related to length of gestation and fetal growth, https://opengwas.io/datasets/finn-b-P16_DISORD_RELATED_LENGTH_GESTATION_FETAL_GROWTH; Haemorrhage in early pregnancy, https://opengwas.io/datasets/finn-b-O15_HAEMORRH_EARLY_PREG#; Pregnancy with abortive outcome, https://opengwas.io/datasets/finn-b-O15_PREG_ABORT; Ectopic pregnancy, https://opengwas.io/datasets/finn-b-O15_PREG_ECTOP; Puerperal sepsis, https://opengwas.io/datasets/finn-b-O15_PUERP_SEPSIS; Other puerperal infections, https://opengwas.io/datasets/finn-b-O15_PUERP_INFECT_OTHER

## Author Contributions

JC, XJ and DB designed the study. RW and BB collected the data. JC and RW performed the MR analysis. JB, ST and JC searched the literature and wrote the manuscript. JC, XJ and DB were responsible for the supervision of the work. All authors contributed to the article and approved the submitted version.

## Acknowledgments

We want to acknowledge the participants and investigators of the MRC Integrative Epidemiology Unit (IEU), FinnGen study and the MiBioGen consortium.

## Publisher’s note

All claims expressed in this article are solely those of the authors and do not necessarily represent those of their affiliated organizations, or those of the publisher, the editors and the reviewers. Any product that may be evaluated in this article, or claim that may be made by its manufacturer, is not guaranteed or endorsed by the publisher.

